# Predictive Modeling of COVID-19 Variant Peak Prevalence and Duration Using GISAID Data Across 15 Countries

**DOI:** 10.64898/2026.02.04.26345559

**Authors:** Yifan Zhang, Rob Paton, Kun Chen, Christopher E. Overton, Jaehun Jung, Youngji Jo

**Author notes:** Corresponding authors Youngji Jo, Jaehun Jung.

## Abstract

**Background:** Rapid emergence and replacement of SARS-CoV-2 variants underscore the need for early and reliable indicators of variant dominance to guide timely public health response. However, early genomic trajectories are typically short, sparse, and noisy, with strong fluctuations and substantial cross-country heterogeneity in sequencing intensity and reporting.

**Methods:** We develop a scalable forecasting framework that predicts whether new variants will reach high prevalence and how long they will persist based on their initial genomic growth patterns. Using more than nine million sequences from 15 countries (GISAID, 2020–2024), we characterize dominance through peak prevalence and duration above 10% and extract early growth descriptors from the first 2–4 weeks after a lineage surpasses 1% frequency. Outcomes were classified using multiple models (GLM, GAM, SVM, CART, Elastic Net, and SuperLearner). We evaluated performance based on accuracy and utilized SHAP analysis to interpret feature importance.

**Results:** The Super Learner ensemble model achieved the best performance, achieving up to 0.76 accuracy for peak-share prediction, and up to 0.70 accuracy for duration classification—substantially outperforming all individual models. SHAP analysis showed that variants achieving high peaks exhibit strong but structurally coherent early growth, whereas prolonged dominance is associated not with early surges but with sustained, moderate short-term fluctuations embedded within a stable trajectory.

**Conclusion:** This framework defines minimum surveillance thresholds (≥100 sequences in 30 days, ≥1% detection share), variant grouping rules, and noise-filtering protocols, enabling cross-country comparison and country-specific forecasting. It provides a lightweight, reproducible early-warning tool for genomic surveillance and real-time epidemic intelligence.

**Significance:** Identifying emerging SARS-CoV-2 variants capable of driving new surges is critical for global preparedness but remains challenging due to sparse early data. We present a machine learning framework that forecasts variant dominance using only the first 2–4 weeks of genomic growth. Analyzing nine million sequences across 15 countries, we reveal two distinct epidemiological signatures: high peak prevalence is driven by explosive, coherent early expansion, while long-term persistence is predicted by sustained, moderate fluctuations rather than initial speed. By establishing minimum surveillance thresholds, this work delivers a scalable, data-efficient early-warning tool that links early genomic signatures of viral fitness to downstream population-level dominance, achieving high predictive accuracy with a minimal number of sequences.

## Introduction

The SARS-CoV-2 pandemic has provided a unique opportunity to observe viral evolution and competition in real time, offering insights into broader respiratory pathogen dynamics [1]. As variants have demonstrated clear patterns of emergence, dominance, and replacement, this viral succession has created a natural experiment in pathogen evolution. Identifying early indicators of dominance carries profound implications for public health planning, including vaccination strategies and resource allocation. Consequently, global health organizations like the Centers for Disease Control and Prevention (CDC) and the World Health Organization (WHO) have emphasized that SARS-CoV-2 variant surveillance remains critical for pandemic preparedness [2], [3].

Despite the global experience of multiple SARS-CoV-2 infection waves, timely identification of variants capable of driving new surges remains difficult due to several intertwined factors. Capturing early signals of variant dominance is particularly challenging because early trajectories are short, sparse, and noisy, with strong daily fluctuations and large cross-country variation in sequencing practices. Defining the initial observation period is also delicate including too many early zeros or stochastic spikes can distort growth signals and obscure true trends. Moreover, the virus’s high mutation rate continually generates a diverse pool of variants with distinct genetic profiles that affect transmissibility, immune escape, and pathogenicity, complicating efforts to identify which low-prevalence lineages (e.g., 0.01–3%) are likely to dominate. The challenge is compounded by variability in sequencing capacity across countries, with low-resource settings often facing delays in generating sufficient genomic data to detect emerging variants early. For instance, while high-sequencing settings (∼1,000 sequences/week) enable robust monitoring, regions with limited infrastructure (∼100 sequences/week) often face detection delays and incomplete genomic coverage. Recent analyses show that 78% of high-income countries sequenced over 0.5% of their COVID-19 cases, compared with only 42% of low- and middle-income countries [4].

Recent modeling studies have made important progress in forecasting SARS-CoV-2 variant dynamics, yet most require data volumes or structures that limit early, global applicability.[5][6] Obermeyer et al. (2022) developed PyR□, a hierarchical Bayesian model that infers lineage fitness by decomposing growth rates into mutation-level effects, enabling identification of amino acid changes associated with increased transmissibility independent of geography or time [7]. While powerful for elucidating molecular drivers of fitness across millions of sequences, PyR□ is computationally intensive and depends on dense, globally distributed data to reliably disentangle mutation effects. In practice, it performs best only after variants reach ≥1% global frequency (roughly corresponding to 100–500 sequences in multiple countries ≥3 consecutive weeks). —and assumes mutation effects are static across space and time, an assumption that may not hold under evolving immunity landscapes and intervention regimes. On the other hand, frequency-based statistical models—such as the multinomial logistic regression frameworks evaluated by Abousamra et al. [8] —can produce accurate short-term forecasts using count data alone. Yet, their reliability deteriorates sharply in settings with sparse or delayed reporting, with analyses suggesting that approximately 1,000 sequences per week are necessary to maintain robust forecast accuracy—a threshold many regions fail to meet. Together, these studies indicate that early variant-frequency trajectories do encode predictive signal, but that forecast reliability is constrained less by algorithmic sophistication than by data availability, timeliness, and calibration. This gap motivates our data-efficient, early-growth–based framework, which links early genomic signatures of viral fitness to downstream population-level dominance, achieving high predictive accuracy with a minimal number of sequences.

Based on comprehensive genomic data from the Global Initiative on Sharing All Influenza Data (GISAID), we analyze lineage dynamics across 15 high-burden countries from January 2020 to May 2024, addressing two fundamental questions: which variants emerge to dominance and how long they sustain it. Leveraging this large dataset, we established minimum surveillance thresholds, variant grouping criteria, noise-filtering protocols, and cross-country comparability, delivering a lightweight and scalable early-warning tool for variant dominance prediction, defined as: weekly peak share and duration of circulation above 10% share. These findings not only advance methodological approaches for genomic surveillance but also provide a reference framework for anticipating the emergence and persistence of future SARS-CoV-2 variants.

## Methods

### Data source

Genomic data for this study were obtained from GISAID (https://www.gisaid.org/) [9]. Established in 2017 as the Global Initiative on Sharing All Influenza Data and later expanded with its EpiCoV repository to include SARS-CoV-2 sequences, GISAID now provides access to millions of viral genomes complete with collection dates, geographic origins and sequencing-platform metadata—enabling researchers worldwide to track the virus’s evolution, transmission, and mutations. Since its launch in January 2020, GISAID has been regularly updated with submissions from various countries, showcasing collaborative efforts in combating the pandemic. Within this shared framework, viral evolution is tracked through the Pango lineage system, in which lineages are defined when clusters of sequences exhibit consistent genetic divergence and epidemiological relevance. Early variants were labeled with letters for major clades (A, B) and numbered sequentially for descendant lineages, providing a standardized and scalable naming structure. Following the WHO/CDC-endorsed Pango lineage nomenclature, the classified lineages collectively comprise 14.3 million publicly available SARS-CoV-2 sequences reported across 223 countries as of September 9, 2024. To ensure broad and representative genomic coverage, we restricted our analysis to the 15 countries with the largest cumulative numbers of reported sequences, yielding a final dataset of 8.2 million sequences spanning 3,680 lineages, collected between December 2019 and September 2024. Among these countries, the United States contributed the largest number of sequences (4.6 million), whereas Austria contributed the fewest (0.2 million). Table 1 summarizes key descriptive statistics for the United States, the United Kingdom, and South Korea, including annual sequencing totals and the five most prevalent variants with their corresponding yearly proportions.

**Table 1.** Descriptive summary of GASID data showing the distribution of raw PANGO lineages in the United States, the United Kingdom, and South Korea.

| Country | Year | Number of Cases | Top Five Pango Lineages (Annual Proportional Share) |
| --- | --- | --- | --- |
| United States | 2020 | 18,903,169 | B.1 (1.1%); B.1.1 (1.1%); B.1.595 (1.1%); B.1.369 (1.1%); B.1.243 (1%) |
|  | 2021 | 196,861,939 | B.1 (1.0%); B.1.617.2 (1%); AY.44 (0.9%); B (0.9%); AY.25 (0.9%) |
|  | 2022 | 227,680,907 | BA.2 (0.7%); BA.1.1 (0.6%); BA.2.12.1 (0.6%); BA.2.3 (0.6%); BA.2.9 (0.5%) |
|  | 2023 | 71,328,981 | XBB.1.5 (0.5%); XBB.1.5.10 (0.4%); XBB.1 (0.4%); XBB.1.5.15 (0.4%); XBB.1.5.49 (0.4%) |
|  | 2024 | 9,457,427 | JN.1 (1.1%); JN.1.8 (1.1%); JN.1.18 (1%); JN.1.7 (1%); JN.1.4 (1.0%) |
| United Kingdom | 2020 | 13,728,198 | B.1.1 (1.2%); B.1 (1.2%); B.1.1.37 (1%); B.1.1.1 (1%); B.1.177 (0.9%) |
|  | 2021 | 84,393,505 | AY.4 (1.5%); B.1.617.2 (1.5%); AY.122 (1.5%); AY.5 (1.5%); AY.111 (1.5%) |
|  | 2022 | 79,974,012 | BA.2 (1.3%); BA.2.3 (1.2%); BA.1.1 (1.2%); BA.2.10 (1.2%); BA.1 (1.2%) |
|  | 2023 | 8,416,483 | XBB.1.5 (0.9%); XBB.1.9.1 (0.7%); CH.1.1.1 (0.7%); XBB.1.5.7 (0.7%); XBB.1.5.18 (0.7%) |
|  | 2024 | 934,727 | JN.1 (2%); JN.1.7 (1.9%); KP.2 (1.7%); KP.3 (1.6%); JN.1.8 (1.5%) |
| South Korea | 2020 | 5,878 | B.1.497 (37.3%); B.1 (9.8%); B.1.1 (8.0%); B.41 (2.9%); A (2.8%) |
|  | 2021 | 215,082 | AY.69 (11.6%); B.1.617.2 (11.6%); AY.122 (9.4%); AY.122.5 (8.0%); B.1.619.1 (6.4%) |
|  | 2022 | 2,827,981 | BA.5.2 (2.0%); BA.5.2.1 (2.0%); BA.5.1 (1.8%); BA.2.3 (1.8%); BA.2 (1.7%) |
|  | 2023 | 4,438,592 | XBB.1.5 (1.2%); XBB.1.16 (1.0%); EG.1.6 (1.0%); EG.1 (1.0%); XBB.1.9.1 (1.0%) |
|  | 2024 | 225,726 | JN.1.4 (4.2%); JN.1 (4.1%); JN.1.20 (3.9%); JN.1.4.5 (3.8%); JN.1.5 (3.4%) |

### Data processing

To ensure the reliability of national-level share estimates, we identified valid analytical periods for each country using sequencing coverage standards recommended by the World Health Organization [10]. A valid period was defined as a continuous stretch of at least three calendar weeks, each containing at least 100 sequences and at least four days with non-zero sampling. This criterion was designed to mitigate the substantial noise and bias introduced by sparse or irregular sampling, which has been shown to distort early growth estimates and peak prevalence inference, particularly in later pandemic phases characterized by slower, noisier, and more heterogeneous variant dynamics [11], [12], [13]. To reliably characterize fitness using early trajectory features, we retained only variant–country combinations fully observed within valid analytical periods. This criterion preserved 24,286 of 26,152 lineages (93%), ensuring that growth and peak prevalence estimates reflect consistent surveillance data rather than artifacts of intermittent sampling. The overall data processing workflow, including data cleaning, phase-based variant grouping, analytical-period selection, and model data generation, is summarized in Figure 1.

**Figure 1.**
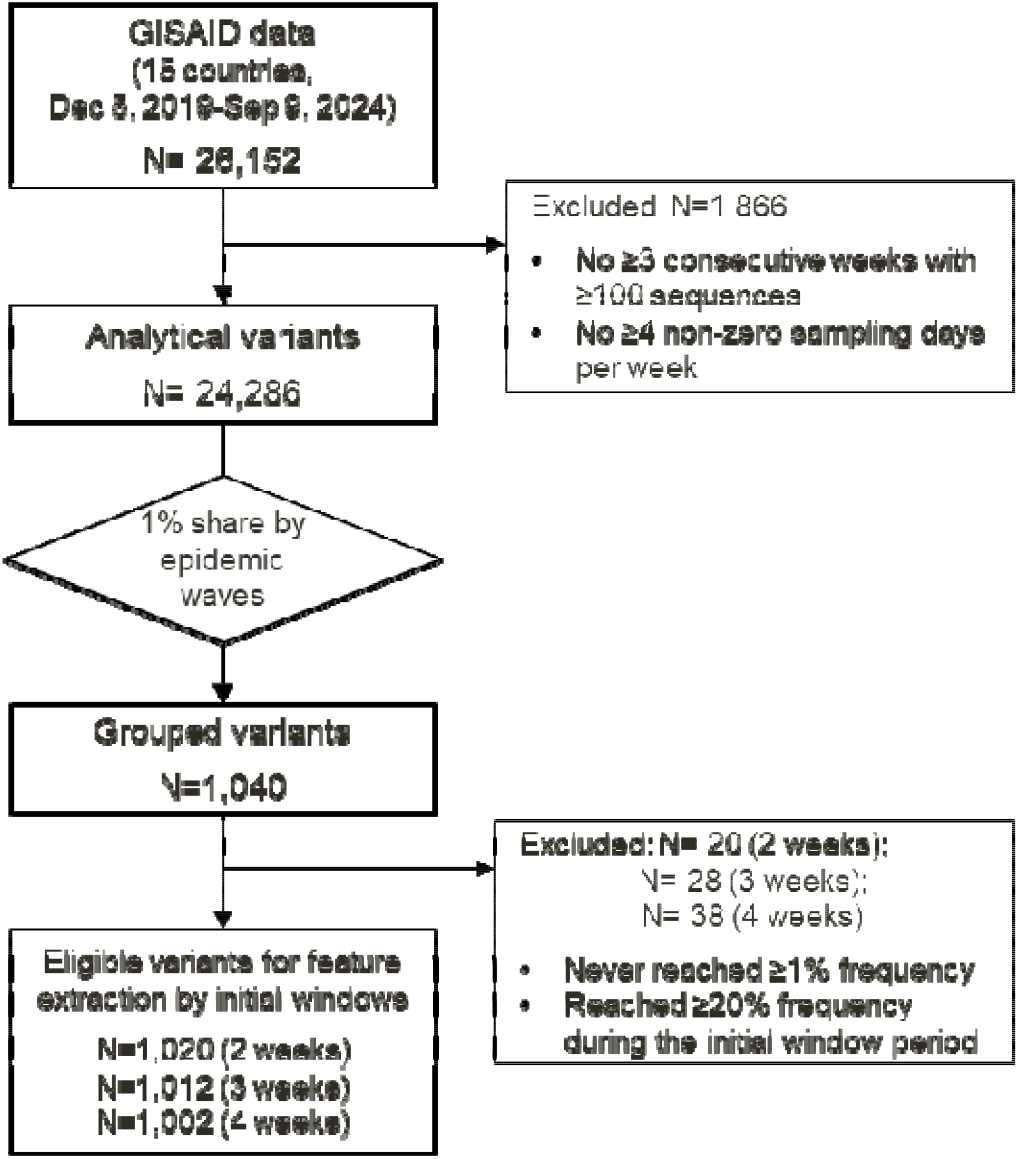
Overview of the data processing workflow, including data inclusion, exclusion, and grouping steps.

### Variants grouping

We segmented the global epidemic into six lineage-specific periods aligned with WHO-recognized phases: pre-VOC (Dec 2019–Sep 2020), Alpha (Oct 2020–May 2021), Delta (Jun–Nov 2021), early Omicron BA.1/BA.2 (Dec 2021–May 2022), mid-Omicron BA.4/BA.5 (Jun–Dec 2022), and late Omicron XBB/JN (Jan 2023– Sep 2024). Within each period, we grouped SARS-CoV-2 lineages into variants using country- and period-specific aggregated case proportions rather than a single global threshold, allowing for substantial cross-country and temporal heterogeneity in genomic surveillance and competitive dynamics. This approach reflects a fundamental shift in selective pressures over time. Early waves were dominated by transmissibility-driven selective sweeps, producing relatively consistent growth and replacement patterns across settings. In contrast, the Omicron era has been shaped by high and heterogeneous background immunity, frequent co-circulation of closely related lineages, and more constrained and variable peak shares. Applying a uniform global grouping rule across all countries and phases would conflate these distinct competitive regimes and systematically under-identify epidemiologically meaningful variants in later periods, where success often appears as moderate but sustained circulation rather than near-complete replacement.

Variant grouping was performed independently within each period. To obtain a global set of grouped variants across the whole study horizon, variant identities were subsequently aggregated across periods based on their unique lineage composition, such that each grouped variant was counted only once globally, even if it met the 1% threshold in multiple periods or countries. Across the six periods, this procedure yielded 1,040 distinct grouped variants, while all remaining low-frequency lineages were collectively assigned to the “others” category within each period. This classification consolidates highly granular lineage data into a tractable number of epidemiologically meaningful groups. Figure 2 illustrates the evolving landscape of grouped SARS-CoV-2 variants in the United States, the United Kingdom, and South Korea.

**Figure 2.**
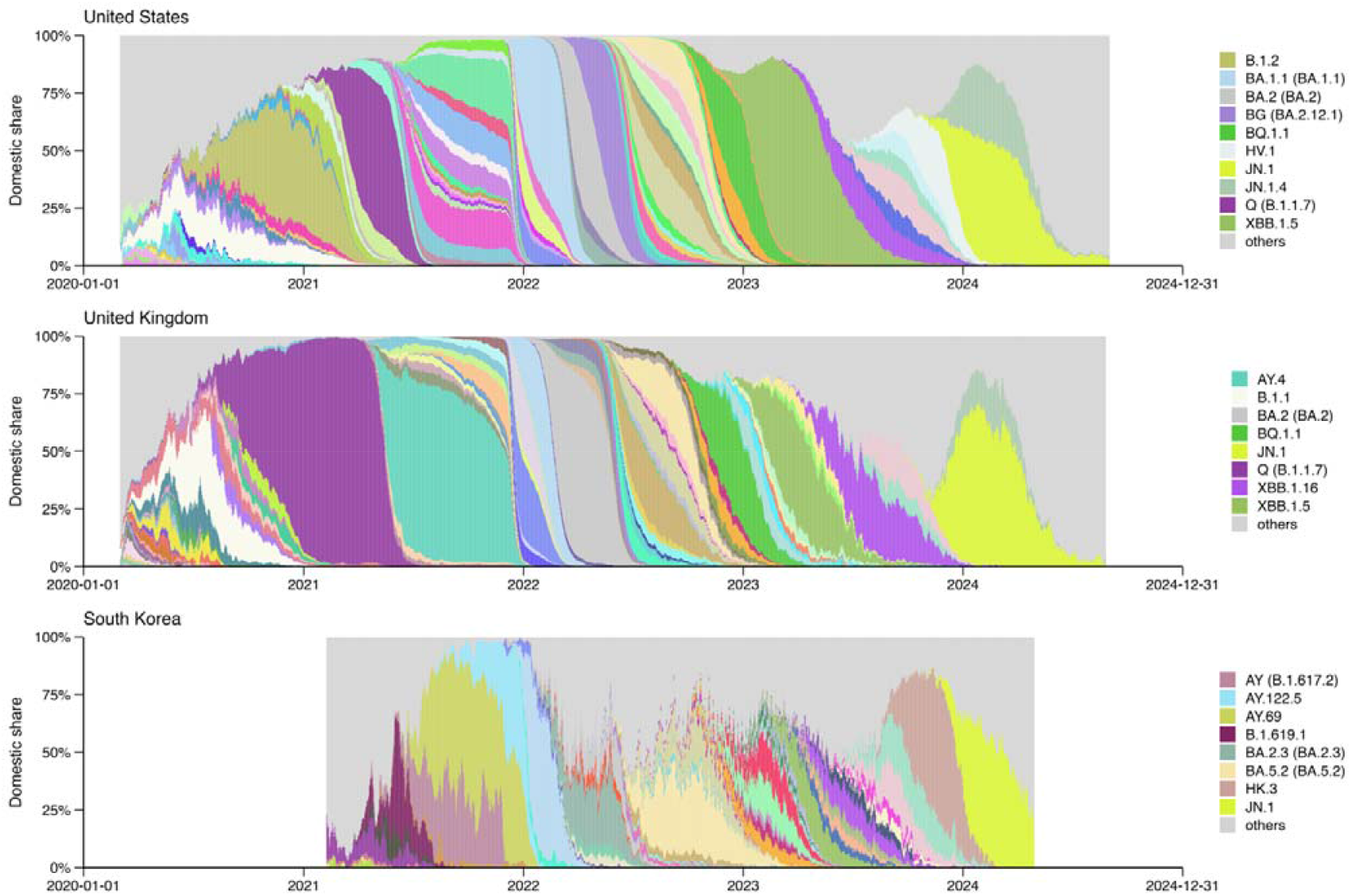
Time-series distribution of grouped SARS-CoV-2 PANGO lineages during the analytical period in the United Kingdom, the United States, and South Korea. Only lineages with weekly maximum share > 20% and > 10% share sustained for > 100 days are shown in the legend.

**Figure 3.**
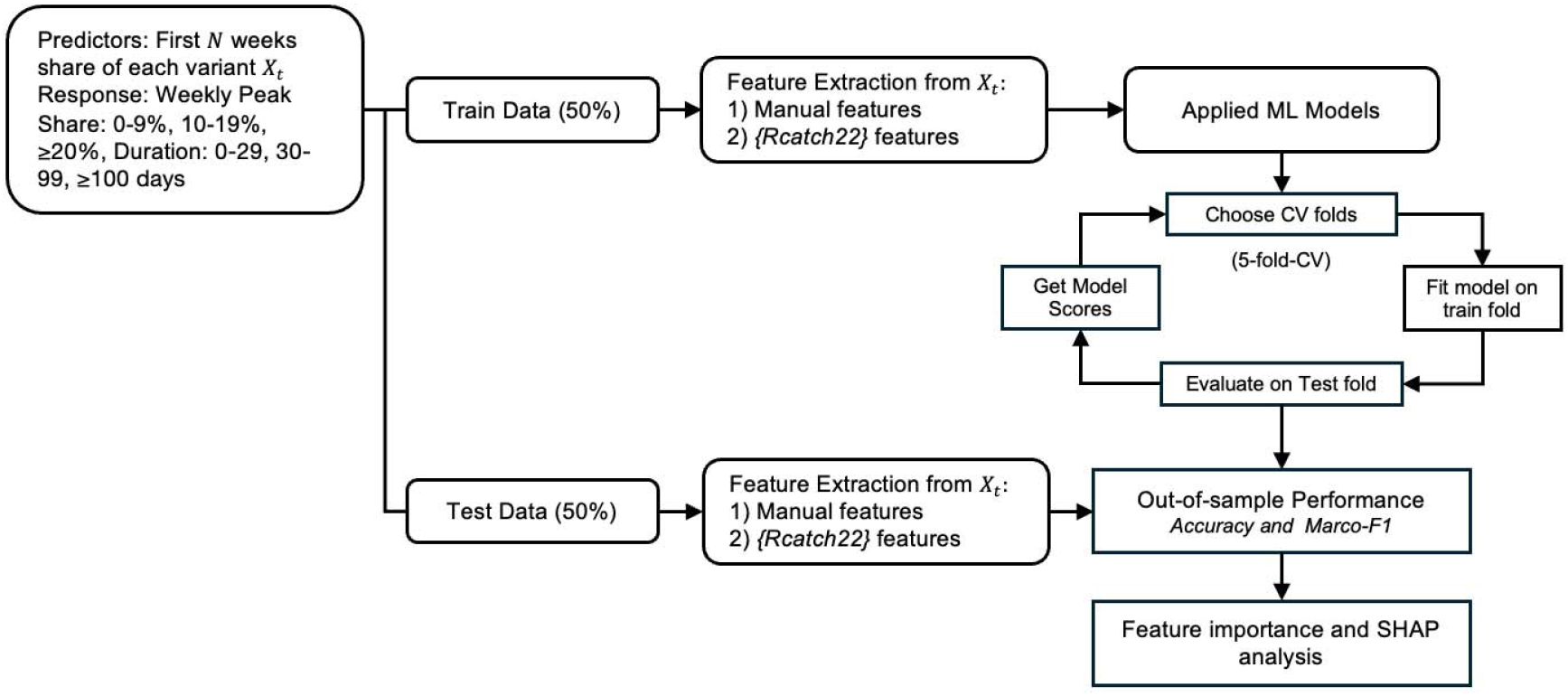
Modeling framework. Variant shares from the first N weeks were used to predict the weekly peak share and duration above 10% share. Features included handcrafted and Rcatch22 descriptors.

### Outcome measures

We examine whether early growth patterns of SARS-CoV-2 variants—captured by their initial national-level share trajectories—forecast two outcomes: the weekly maximum domestic share (peak) and the number of days (duration) a variant maintains a domestic share above 10% [14]. Both outcomes are discretized into categorical levels for the primary analyses to facilitate robust comparison across variants and countries. These two measures better capture a variant’s competitive strength and epidemiological relevance than cumulative share–based metrics, which can be inflated by prolonged low-level circulation. Daily sequence counts were aggregated using a rolling seven-day window, within which the variant’s proportion was computed as its sequence count divided by the total sequences collected over the same window. This windowed formulation reduces sensitivity to day-level reporting noise while capturing the variant’s strongest observed competitive position[15]. This continuous measure, ranging from 0 to 1, characterizes the intensity of a variant’s competitive advantage. Its empirical distribution is highly right-skewed, with most variants attaining only modest peaks (median ≈ 0.08; IQR: 0.03–0.20). To mitigate the influence of extreme values and enhance model stability, we discretized peak share into an ordered three-level outcome: 0-9%, 10– 19%, and ≥20%. The category thresholds were informed by the empirical distribution of peak share, with cut points guided by the observed interquartile range. A >20% domestic share was considered dominant, as high background immunity and multi-lineage competition in the post-Omicron era constrain peak prevalence, making such levels indicative of a clear competitive advantage. Given that values exceeding 20% fall within the extreme upper tail of the distribution, this uppermost category is interpreted as representing high-peak, or dominant, variants. Similarly, the empirical distribution of duration is highly skewed, with nearly one-third of variants never exceeding the 10% threshold for more than 30 days. Using duration as a continuous outcome would therefore place disproportionate weight on a small number of long-lasting variants and yield unstable estimates for the majority of low-duration observations. To reduce sensitivity to extreme values while preserving epidemiologically meaningful distinctions in persistence, duration was discretized into three ordinal categories: 0–29, 30–99, and ≥100 days [16].

### Predictive features

The start point of the initial period was defined as the first time the variants reached a 1% share in their respective countries. This threshold was selected to mark a reproducible surveillance-relevant onset, at which variant detection is sufficiently stable to support reliable estimation of early growth dynamics. From this point, we considered a different set of observation periods from 14 to 28 days. From these early share sequences, we then constructed two complementary sets of predictors. The first group comprised handcrafted, epidemiologically motivated predictors designed to capture basic growth dynamics. These included the overall log-linear slope (overall growth or decline rate), short-window slopes over seven days, the longest increasing run (consecutive growth days), and the non-zero count (number of days with non-zero share). All features were computed from early-stage share trajectories using base R functions and custom routines. The second set was drawn from the *catch22* library [17], accessed via the *Rcatch22* package, which provides 22 minimally redundant descriptors of time-series structure such as entropy, fluctuation scaling, and motif frequency. The catch22 features are computationally efficient, robust to short and noisy sequences, and broadly applicable without parameter tuning, which can capture the heterogeneity of early trajectories. By combining interpretable, domain-informed measures with standardized machine-learning–oriented descriptors, this approach balances epidemiological interpretability with statistical power and enhances generalizability across diverse variant profiles. Together, these features quantify each trajectory’s level, variability, early acceleration, and persistence, providing a rich basis for downstream prediction of peak share and time to dominance. The specific feature set was derived following established frameworks for interpretable time-series feature extraction [16] and prior applications to SARS-CoV-2 variant growth [13].

### Prediction model

We employed a diverse set of machine learning regressors suited for heterogeneous temporal data. These included generalized linear models [18], generalized additive models [19], regularized methods such as Elastic Net [20], kernel-based models [21], neural networks [22], and tree-based approaches [23]. We further applied the SuperLearner R package [24], an ensemble learning framework that optimally combines multiple algorithms. Specifically, we applied a SuperLearner ensemble to balance interpretability, flexibility, and regularization, which pools complementary models—CART for nonlinear patterns, GLM for main effects, and Elastic Net for correlated predictors—through cross-validated stacking to approximate the optimal weighted combination. By jointly borrowing strength across countries and feature sets, the approach stabilizes variant fitness estimates and reduces noise in data-sparse settings. Variant share trajectories are inherently dependent, as they are derived from relative proportions and observed longitudinally over time. This dependence is explicitly acknowledged and is compatible with the predictive focus of the analysis, which does not rely on independence assumptions required for formal statistical insference. Figure 2 illustrates the modeling workflow. Variants were randomly partitioned into training (50%) and test (50%) sets across all countries and epidemic periods. Model hyperparameters were optimized within the training set using five-fold cross-validation, with fold-averaged accuracy and macro-F1 used as selection criteria [25]. Final model performance was evaluated on the independent test set, which was not involved in model fitting or tuning.

### Evaluation of model performance and interpretation

For the categorical outcomes of weekly peak share and dominance duration, we report out-of-sample overall accuracy and macro-F1, evaluated on the held-out test set. Accuracy summarizes the proportion of correctly classified variants, while macro-F1 equally weights precision and recall across classes to account for class imbalance [26]. Classification decision thresholds and model hyperparameters were selected within the training set via cross-validation and then fixed for evaluation on the test set. To further interpret model behavior, we applied variable importance analysis and SHAP decomposition [27], which together identify key predictors and quantify their contributions at both global and local levels. The entire workflow, from feature extraction to model fitting, evaluation, and interpretation, is summarized in Figure 2.

## Results

### Comparison of model prediction and observed dominant variants

Table 3 summarizes model performance for the three most well-sampled countries under the operational definition of dominance (peak >20% and duration ≥100 days). In both the United States and the United Kingdom, the model showed strong consistency: nearly all major lineages— including BA.2, BA.5 sublineages, BQ.1.1, XBB.1.5, and JN.1—were correctly classified for both peak and duration, with only early variants (e.g., B.1.2 in the U.S., B.1.1 and B.1.1.7 in the U.K.) showing underestimation of peak magnitude. Results in South Korea were more heterogeneous: dominant variants were generally detected, but peak share was frequently underpredicted for Delta-related AY lineages and for later Omicron sublineages such as JN.1. Despite this, duration classification remained highly accurate across all three countries. Overall, the model achieves high agreement in the U.S. and U.K., with reduced peak-prediction stability in South Korea due to more variable early growth dynamics.

**Table 3.** Comparison of observed and predicted variant dominance in selected countries, defined as duration ≥100 days and peak >20%.

| Country | Lineage | Onset date | Offset date | Pred. Peak | Obs. Peak | Peak. Acc. | Pred. Dur. | Obs. Dur. | Dur. Acc. |
| --- | --- | --- | --- | --- | --- | --- | --- | --- | --- |
| United States | B.1.2 | 2020-03-09 | 2023-07-17 | 0.10–0.20 | $>0.20$ | F | 31–100 | 100+ | F |
| | Q (B.1.1.7) | 2020-03-12 | 2024-03-04 | 0.10–0.20 | $>0.20$ | F | 31–100 | 100+ | F |
| | BA.2 (BA.2) | 2020-04-03 | 2024-08-22 | $>0.20$ | $>0.20$ | T | 100+ | 100+ | T |
| | XBB.1.5 | 2020-04-04 | 2024-07-16 | $>0.20$ | $>0.20$ | T | 100+ | 100+ | T |
| | AY.103 | 2020-05-14 | 2023-02-01 | $>0.20$ | $>0.20$ | T | 100+ | 100+ | T |
| | BG (BA.2.12.1) | 2020-08-18 | 2024-08-28 | $>0.20$ | $>0.20$ | T | 100+ | 100+ | T |
| | BF (BA.5.2.1) | 2021-11-05 | 2024-04-11 | $>0.20$ | $>0.20$ | T | 100+ | 100+ | T |
| | BQ.1.1 | 2021-11-30 | 2024-03-27 | $>0.20$ | $>0.20$ | T | 100+ | 100+ | T |
| | HV.1 | 2023-01-09 | 2024-08-16 | $>0.20$ | $>0.20$ | T | 100+ | 100+ | T |
| | JN.1 | 2023-01-09 | 2024-08-31 | $>0.20$ | $>0.20$ | T | 100+ | 100+ | T |
| | JN.1.4 | 2023-09-13 | 2024-08-29 | $>0.20$ | $>0.20$ | T | 100+ | 100+ | T |
|  | others (n = 68) | 2020-03-02 | 2024-08-16 | - | - | - | - | - | - |
| United Kingdom | B.1.1 | 2020-03-02 | 2022-06-23 | 0.10–0.20 | $>0.20$ | F | 31–100 | 100+ | F |
| | Q (B.1.1.7) | 2020-03-03 | 2022-03-03 | 0.10–0.20 | $>0.20$ | F | 100+ | 100+ | T |
| | BA.2 (BA.2) | 2021-01-01 | 2024-01-13 | $>0.20$ | $>0.20$ | T | 100+ | 100+ | T |
| | BA.5.1 (BA.5.1) | 2022-01-12 | 2024-05-10 | $>0.20$ | $>0.20$ | T | 100+ | 100+ | T |
| | BA.5.2 (BA.5.2) | 2022-01-17 | 2024-02-12 | $>0.20$ | $>0.20$ | T | 100+ | 100+ | T |
| | BQ.1.1 | 2022-02-11 | 2024-01-01 | $>0.20$ | $>0.20$ | T | 100+ | 100+ | T |
| | XBB.1.5 | 2022-10-30 | 2024-05-28 | 0.10–0.20 | $>0.20$ | F | 100+ | 100+ | T |
| | XBB.1.16 | 2023-01-01 | 2024-04-17 | $>0.20$ | $>0.20$ | T | 100+ | 100+ | T |
| | JN.1.4 | 2023-09-18 | 2024-08-25 | $>0.20$ | $>0.20$ | T | 100+ | 100+ | T |
| | JN.1 | 2023-09-21 | 2024-08-24 | $>0.20$ | $>0.20$ | T | 100+ | 100+ | T |
|  | others (n = 76) | 2020-03-02 | 2024-07-10 | - | - | - | - | - | - |
| South Korea | B.1.619.1 | 2021-02-22 | 2021-09-21 | $>0.20$ | $>0.20$ | T | 100+ | 100+ | T |
| | AY (B.1.617.2) | 2021-04-22 | 2023-07-11 | 0.10–0.20 | $>0.20$ | F | 100+ | 100+ | T |
| | AY.69 | 2021-05-15 | 2022-02-10 | 0.10–0.20 | $>0.20$ | F | 100+ | 100+ | T |
| | AY.122.5 | 2021-08-08 | 2022-03-04 | 0.10–0.20 | $>0.20$ | F | 100+ | 100+ | T |
| | BA.2.3 (BA.2.3) | 2021-12-29 | 2023-03-15 | $>0.20$ | $>0.20$ | T | 100+ | 100+ | T |
| | BF (BA.5.2.1) | 2022-05-10 | 2023-04-15 | 0.10–0.20 | $>0.20$ | F | 31–100 | 100+ | F |
| | BA.5.2 (BA.5.2) | 2022-05-11 | 2024-01-22 | $>0.20$ | $>0.20$ | T | 100+ | 100+ | T |
| | XBB.1.5 | 2022-11-23 | 2023-12-11 | 0.10–0.20 | $>0.20$ | F | 100+ | 100+ | T |
| | EG.5.1.1 | 2023-05-16 | 2024-03-27 | 0.10–0.20 | $>0.20$ | F | 100+ | 100+ | T |
| | HK.3 | 2023-07-04 | 2024-04-15 | $>0.20$ | $>0.20$ | T | 100+ | 100+ | T |
| | JN.1 | 2023-10-24 | 2024-04-28 | 0.10–0.20 | $>0.20$ | F | 31–100 | 100+ | F |
|  | others (n = 37) | 2021-02-08 | 2024-02-20 | - | - | - | - | - | - |

### Country-specific model performance

Figures 4(a) and 4(b) compare classification accuracy across 15 countries using 21-day inputs for predicting peak and duration categories, respectively. Across both tasks, the SuperLearner ensemble shows consistently strong performance across countries, with particularly high accuracy in Denmark, the United Kingdom, Spain, and the United States. For peak prediction, CART attains accuracy comparable to SuperLearner in several countries, while for duration prediction, SVM performs similarly well in selected settings. However, the performance of these individual models varies more across countries and outcomes. In contrast, SuperLearner maintains more stable accuracy across heterogeneous national contexts. Overall, these results suggest that ensemble learning provides a robust and reliable predictive framework for variant growth forecasting across diverse countries and outcome definitions.

**Figure 4(a).**
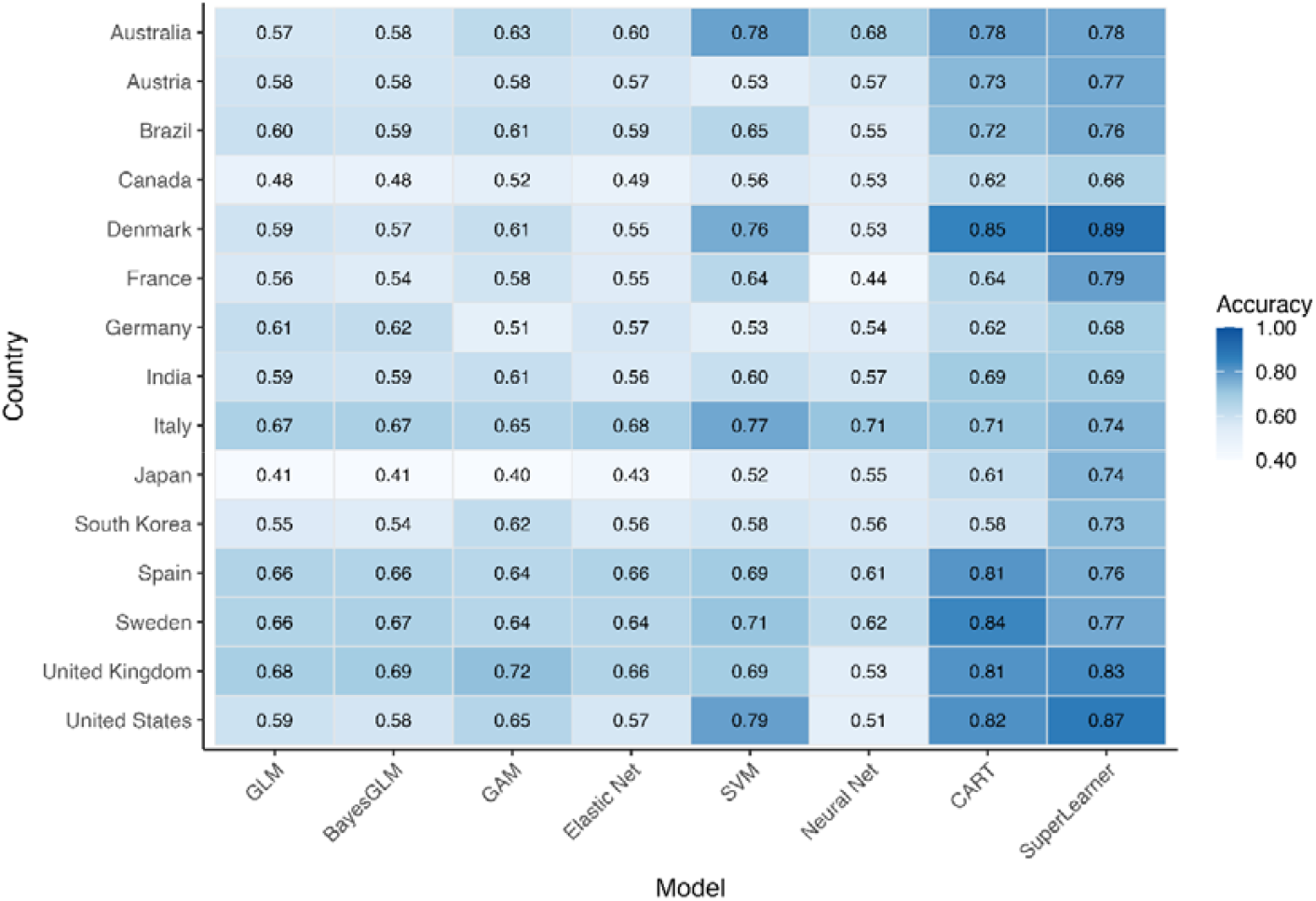
Heatmap of accuracy across models and countries for predicting peak categories using a 21-day input. Blue = higher, white = lower.

**Figure 4(b).**
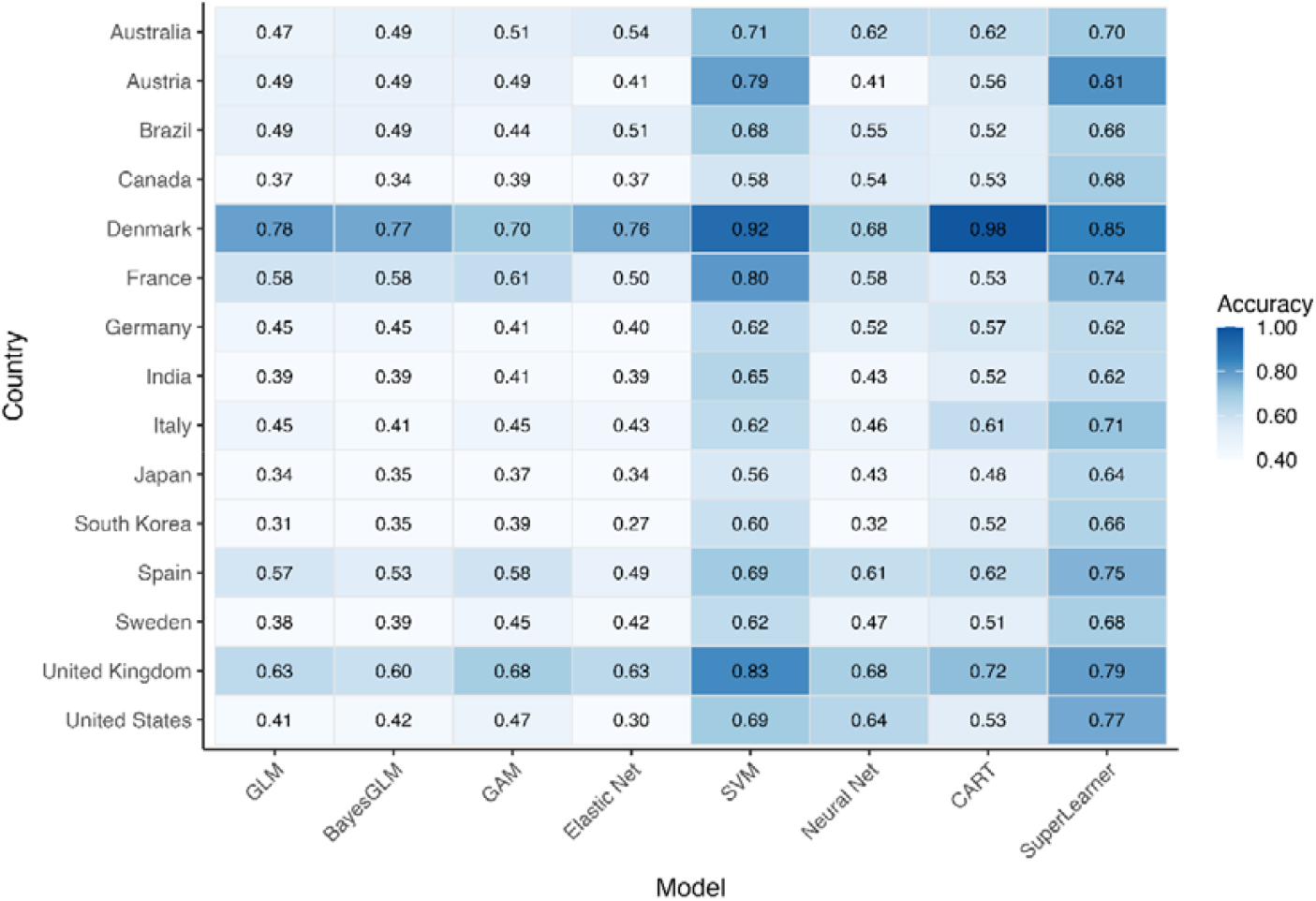
Heatmap of accuracy across models and countries for predicting duration categories using a 21-day input. Blue = higher, white = lower.

**Figure 5.**
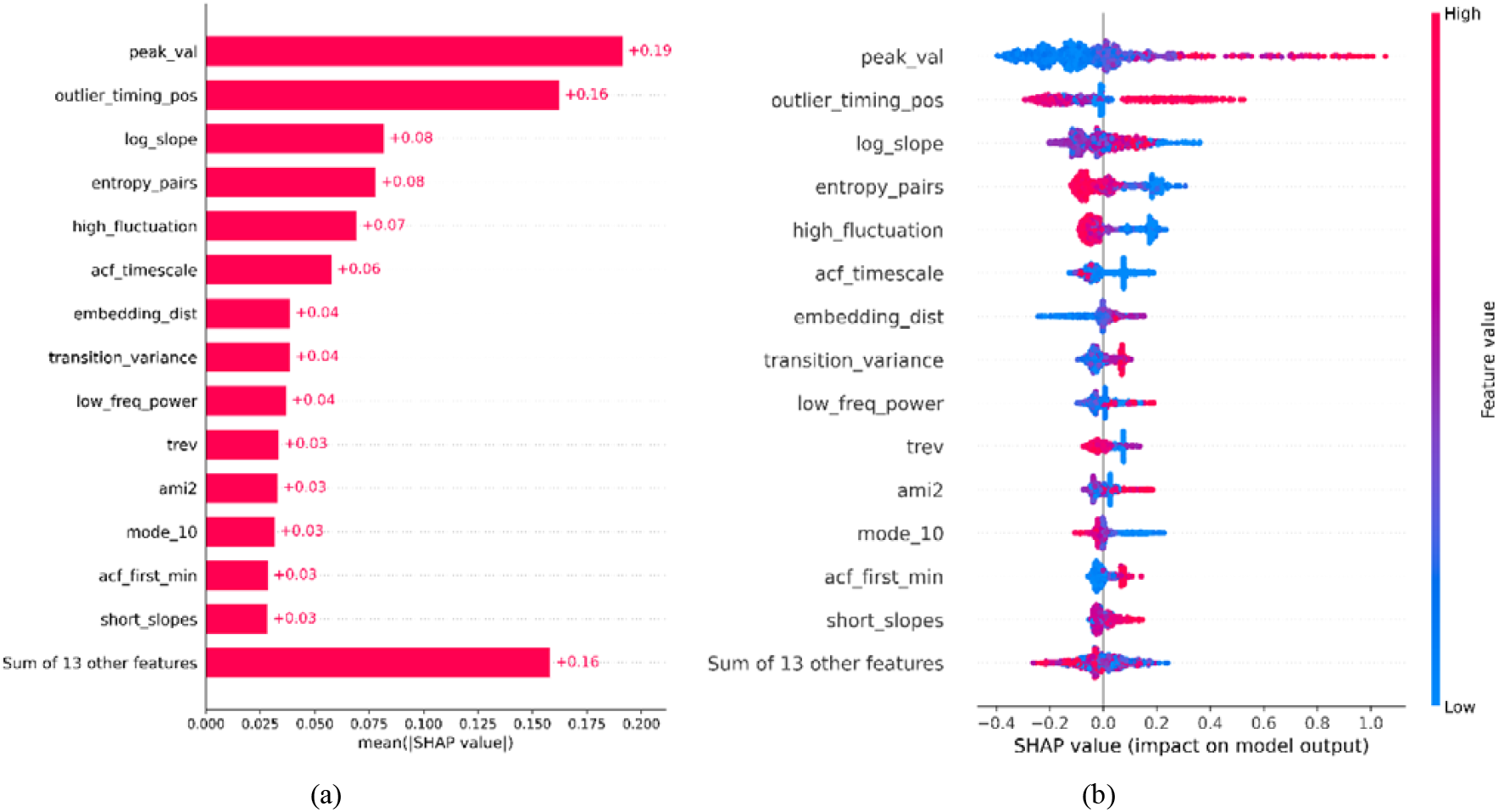
SHAP-based feature attribution for the SuperLearner model (21-day input) predicting variants weekly peak prevalence across 15 countries. (a) Global feature importance based on mean absolute SHAP values. (b) SHAP summary (beeswarm) plot illustrating the distribution and direction of feature effects.

**Figure 6.**
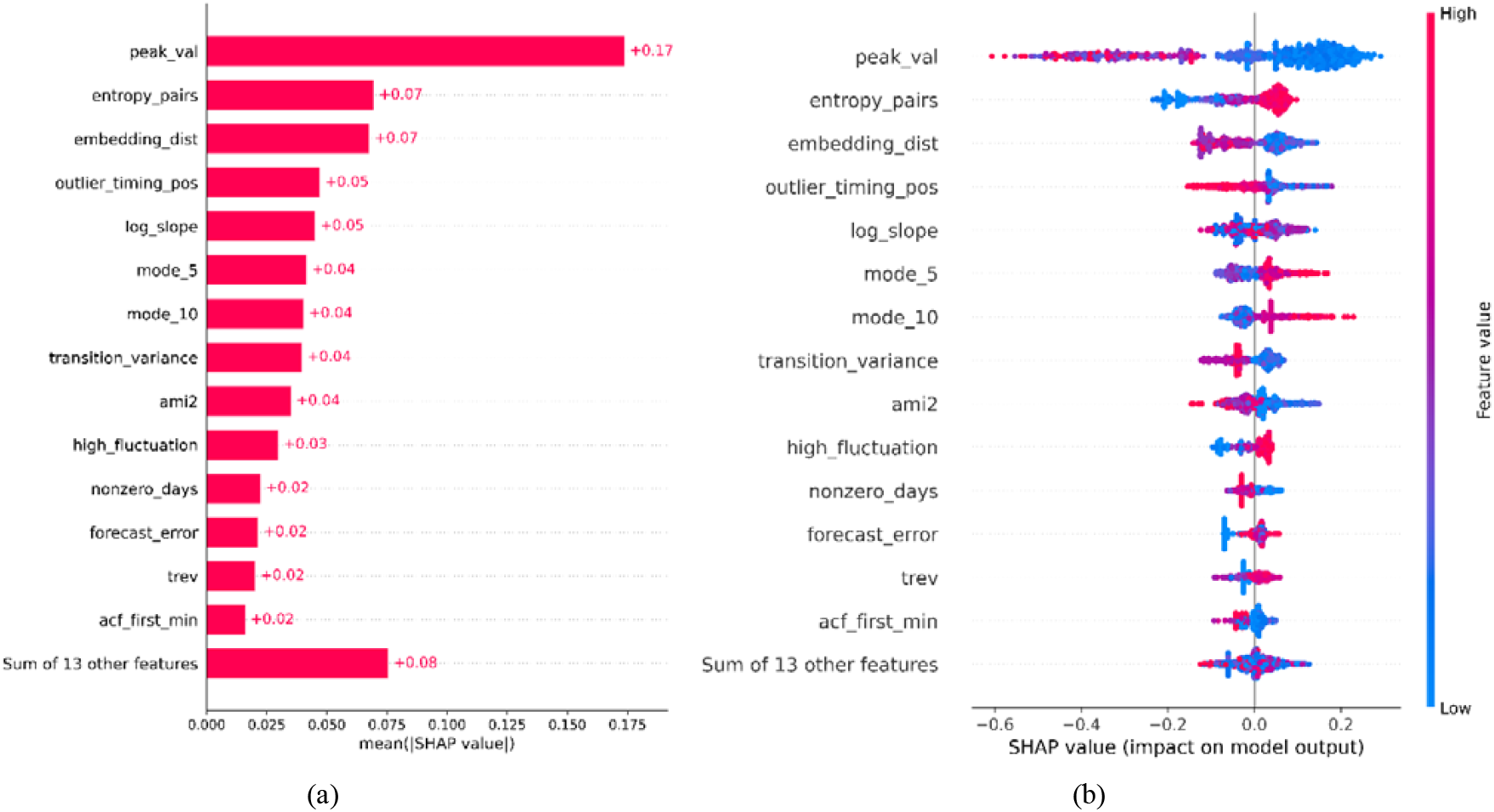
SHAP-based feature attribution for the SuperLearner model (21-day input) predicting variants duration across 15 countries. (a) Global feature importance based on mean absolute SHAP values. (b) SHAP summary (beeswarm) plot illust ating the distribution and direction of feature effects.

### SHAP analysis

The SHAP analysis for the peak-share outcome (>20 percent) shows that both the magnitude and structural coherence of early growth are key determinants of whether a lineage reaches a high weekly peak (Figure 4a). The early maximum value (*peak_val*) exerts the strongest positive influence, indicating that lineages with substantial early expansion are most likely to achieve high peak prevalence. Later positive deviations (*outlier_timing_pos*) further reinforce this signal, while early isolated spikes tend to reduce predicted peaks, reflecting their interpretation as noise rather than evidence of sustained transmission. Structural characteristics of the trajectory refine these effects: higher entropy_pairs consistently lowers predictions, indicating that irregular symbolic patterns are not conducive to strong expansion, whereas longer autocorrelation timescales (*acf_timescale*) and moderate short-term fluctuation (*high_fluctuation*) reflect more persistent and coherent growth. Measures of geometric and nonlinear structure, including *embedding_dist* and *ami2*, similarly distinguish lineages whose early trajectories are organized and dynamically consistent. Overall, variants that combine strong early amplification with stable and structurally coherent growth are more likely to achieve high peak share.

The SHAP analysis for dominance duration (>100 days) reveals a distinct pattern from the peak-share outcome (Figure 4b). Early sharp peaks, reflected in *peak_val*, reduce predicted duration, indicating that rapid initial surges are characteristic of short-lived waves rather than sustained circulation. In contrast, greater short-horizon forecast variability (*forecast_error*) and stronger incremental day-to-day fluctuations (*high_fluctuation*) contribute positively, suggesting that lineages exhibiting recurrent, moderate short-term variation—rather than a single early spike—are more likely to persist. Higher *embedding_dist* and elevated *transition_variance* reduce predicted duration, implying that geometric irregularity and unstable state-switching signal transient rather than resilient growth. Overall, prolonged dominance appears to arise not from explosive early expansion but from continued small-scale dynamism embedded within a coherent and structurally stable growth trajectory.

## Discussion

In this study, we demonstrate that the earliest genomic growth trajectories, captured within just two to four weeks of a dominance. By integrating domain-specific growth indicators with standardized time-series descriptors, our SuperLearner ensemble framework achieved high predictive accuracy for both peak prevalence and duration across diverse national contexts, consistently outperforming individual algorithms. Crucially, our interpretability analysis reveals that these two dimensions of dominance are driven by distinct dynamical signatures: while high peak prevalence is primarily determined by the magnitude and structural coherence of early expansion, prolonged duration is associated not with explosive initial surges, but with sustained, moderate fluctuations embedded within a stable growth trajectory. These findings validate a scalable, sequence-only approach to real-time surveillance, providing public health systems with a lightweight early-warning tool that operates well before variants approach their national peaks.

In our framework, predictive uncertainty primarily stems from two intertwined sources: early growth and detection delay [28]—which together shape the heterogeneity observed across countries [29]. Variants exhibiting extremely rapid initial expansion compress both biological signal and stochastic noise into short time windows, making growth-related features (e.g., short-window slopes, fluctuation metrics) highly sensitive to sampling variability. In high-coverage settings such as Denmark or the United Kingdom, dense sequencing smooths these fluctuations and yields stable slope estimates; in contrast, sporadic sampling in countries like Brazil or South Korea amplifies volatility, occasionally misclassifying transient spikes as sustainable growth. Detection delays further compound these disparities by shifting each country’s analytical “Day 0” away from the true biological onset. In low-throughput systems, a variant may already be mid-growth when it first exceeds the 1 % threshold, leading to truncated or distorted early trajectories and underestimation of eventual dominance. This asymmetry explains why the model achieved near-perfect accuracy in data-rich settings but tended to underpredict peaks and durations in countries with limited sequencing continuity. Naïve growth-rate models struggle under these conditions, as a single spurious point or reporting lag can heavily bias slope estimates. Machine learning approaches are better suited because they integrate multiple features beyond simple growth rates, capture variability and persistence, and learn from diverse historical examples, making them more robust to noise and heterogeneity.

The country-specific results highlight a fundamental shift in the mechanisms governing variant dominance and forecasting uncertainty over the course of the pandemic. In the early phase, when population immunity was low, variant success was largely driven by intrinsic transmissibility, producing rapid, high-signal selective sweeps that yielded relatively consistent early growth patterns and uniform model performance across countries. As global immunity accumulated—through vaccination and prior infection—variant success increasingly depended on immune escape within heterogeneous national immune landscapes, weakening the direct relationship between early growth rate and eventual peak prevalence and amplifying cross-country heterogeneity. Consistent with this transition, Figgins and Bedford (2025)[30] demonstrated that SARS-CoV-2 fitness evolved from transmissibility-driven to immune-escape– driven dynamics, with time-varying selective pressures shaped by population immunity rather than fixed viral traits. Under these conditions, long-term dominance reflects sustained competitiveness under immune pressure rather than explosive early expansion, explaining why duration is more robustly predicted than peak magnitude in later phases. Importantly, the observed heterogeneity across countries reflects this deeper epidemiological shift toward immune-structured, context-dependent persistence, rather than model failure, underscoring the value of early identification of persistent variants over precise peak prediction in the post-Omicron era.

Our findings complement and extend recent molecular-level approaches to forecasting viral evolution, including EVEscape [31] and viral language models. Thadani et al. (2023)[31] demonstrated that pre-pandemic mutational landscapes can anticipate immune escape potential by learning antigenic constraints from deep mutational scanning and structural data, while Hie et al. (2021) [31] showed that protein language models encode evolutionary and functional constraints that predict escape at the mutation level. These frameworks effectively identify the intrinsic potential for immune escape by learning antigenic and evolutionary constraints from pre-pandemic data, they do not directly resolve how those molecular advantages translate into population-level dominance in a crowded fitness landscape. Our results address this gap by showing that early genomic growth trajectories encode two distinct epidemiological dimensions of fitness: peak prevalence, reflecting the strength and coherence of early expansion, and dominance duration, reflecting sustained competitiveness within an immune-structured, multi-lineage environment. Importantly, we find that ‘molecularly fit’ variants do not necessarily achieve prolonged dominance unless their early growth exhibits structural coherence (e.g., moderate sustained fluctuations) rather than just explosive early surges. This distinction is crucial: while molecular models predict the *capacity* for escape, our trajectory-based framework captures the *realization* of that capacity, revealing that long-term persistence in complex immune landscapes depends on stable, structurally organized growth rather than the sheer magnitude of initial expansion. By mapping early sequence-derived growth outcomes—where molecular, immunological, and ecological pressures are already aggregated—to downstream peak and persistence, our framework provides a complementary population-level lens that connects molecular fitness signals to real-time epidemic impact.

A major contribution of this study is its systematic cross-country comparison of variant dynamics across distinct pandemic phases, revealing how the drivers of dominance and uncertainty shifted over time. In the early pandemic, low background immunity produced high signal-to-noise, transmissibility-driven selective sweeps, making early slopes and magnitude-based features highly predictive of both peak and duration. In contrast, the Omicron era reflects a crowded fitness landscape shaped by heterogeneous immunity and intense multi-lineage competition, where early growth signals are weaker, noisier, and structurally more complex. Under these conditions, structural descriptors— such as entropy, autocorrelation, and fluctuation dynamics—become critical, and dominance duration is more reliably predicted than peak magnitude. Consistent with this shift, our analyses show that early growth signatures observed within just 2–4 weeks remain highly informative for forecasting both peak intensity and persistence, even as the biological drivers of success evolve, if surveillance thresholds and observation windows are carefully defined. By establishing practical minimum requirements (e.g., ≥100 sequences in the previous 30 days and a 1% detection threshold) and a standardized early-observation framework, we demonstrate that sequence-only time-series features can support accurate, comparable, and scalable forecasting across diverse epidemiological and surveillance settings.

This study has several methodological and data-related limitations that warrant consideration. First, although the analytical window was anchored to the first observation of a 1% variant share to mitigate stochastic noise in the earliest detections, this design inevitably truncates part of the emergence phase for variants with very low eventual prevalence. Consequently, such lineages may lose subtle but informative early dynamics, limiting the model’s ability to fully characterize weakly competitive or self-limiting variants. Second, while pooling data from 15 countries enhanced statistical power and allowed for a robust cross-national framework, it may also have introduced sampling bias toward countries with dense, continuous sequencing efforts (e.g., the United States, Denmark, and the United Kingdom), potentially underrepresenting signals from regions with sparser genomic surveillance. Third, cross-country heterogeneity in model performance likely reflects structural differences in sequencing intensity, coverage, data continuity, reporting lags, as well as variation in public health interventions and epidemic baselines, which mediate how early growth signals translate into eventual dominance. On average, variants required about four months (116 days) to reach their national peak after first surpassing the 1% threshold, with the longest interval observed in Italy (206 days) and the shortest in Denmark (72 days). In contrast, our model relies on only two to five weeks of early data to generate forecasts—capturing signals long before the peak occurs. This substantial gap highlights the model’s practical value as an early-warning tool, capable of anticipating dominant trajectories well ahead of observed epidemic surges. Future studies could incorporate hierarchical or region-weighted model extensions that account for sampling bias (country-specific detection delays, surveillance cadence) and evolving immunity landscapes [32], which drive much of the cross-country heterogeneity in forecasting performance. Such enhancements would further improve model generalizability and robustness to uneven data quality, supporting the development of lightweight, scalable early-warning systems for global genomic surveillance. Pandemic preparedness will require multi-strategic AI frameworks combining genomic, immunological, and host data to anticipate immune escape, guide vaccine updates, and strengthen early-warning systems—while ensuring ethical governance to mitigate dual-use biosecurity risks [33].

This study introduces a scalable, sequence-only forecasting framework that predicts SARS-CoV-2 variant dominance using just 2–4 weeks of early genomic growth data after a lineage first exceeds 1% prevalence. By integrating handcrafted growth indicators with a standardized set of Rcatch22 time-series descriptors, the approach captures both the magnitude and structural coherence of early expansion. Across 15 countries, the Super Learner ensemble, which combines GLM, Elastic Net, and CART, consistently outperformed individual models, achieving up to 0.85 accuracy for peak-share classification and 0.77 accuracy for dominance-duration prediction. These results demonstrate that reliable forecasts of both peak intensity and persistence can be obtained months before variants reach their national peak, highlighting the utility of early genomic trajectories for real-time epidemic intelligence. Beyond predictive performance, this work establishes a unified framework for genomic surveillance, including minimum sampling thresholds (≥100 sequences within 30 days), a 1% detection benchmark for defining emergence, standardized variant-grouping procedures, and noise-filtering rules that ensure comparability across heterogeneous sequencing environments. SHAP analyses further reveal that variants achieving high peaks are characterized by strong but structurally coherent early growth, whereas longer dominance durations are associated with sustained moderate fluctuations rather than sharp early surges. Together, these methodological and interpretive advances provide a lightweight, reproducible, and globally scalable foundation for early-warning systems capable of anticipating emerging variant trajectories and supporting proactive public health decision-making across diverse resource settings.

## Data Availability

The SARS-CoV-2 genome sequences analyzed in this study were obtained from the GISAID database (https://www.gisaid.org), subject to its data access and usage agreements

## Data and Code Availability

All analysis code and processed data used in this study are publicly available at: https://github.com/UConn-Health-Disease-Modeling/2025_Variants_Prediction.git

## Funding

This research was supported by a grant of the Korea Disease Control and Prevention Agency (KDCA), Republic of Korea (grant number: 2025-03-002) and New Faculty Startup Fund from UConn Health (grant number 208069-10100-72525-10). The funders had no role in study design, data collection and analysis, decision to publish, or preparation of the manuscript.

## Notes

### Competing Interest Statement

The authors have declared no competing interest.

### Funding Statement

This research was supported by the Korea Disease Control and Prevention Agency (KDCA), Republic of Korea (grant number 2025-03-002), and by a New Faculty Startup Fund from UConn Health (grant number 208069-10100-72525-10). The funders had no role in the study design, data collection and analysis, decision to publish, or preparation of the manuscript.

